# Prior Covid-19 and high RBD-IgG levels correlate with protection against VOC-δ SARS-CoV-2 infection in vaccinated nursing home residents

**DOI:** 10.1101/2021.09.21.21263880

**Authors:** Hubert Blain, Edouard Tuaillon, Amandine Pisoni, Laure Soriteau, Elodie Million, Marie-Suzanne Léglise, Isabelle Bussereau, Stéphanie Miot, Yves Rolland, Marie-Christine Picot, Jean Bousquet

**Affiliations:** Department of Internal Medicine and Geriatrics, MUSE University, Montpellier, France; Laboratory of Virology, INSERM U 1058/EFS, University hospital, Montpellier, France; Gérontopôle de Toulouse, INSERM 1027; Toulouse, France; Clinical research and epidemiology unit, University hospital, Montpellier, France; Department of Dermatology and Allergy, Universitätsmedizin, Berlin, Germany and University Hospital, Montpellier, France

## Abstract

**Background:** Nursing Home (NH) residents are at high risk of serious illness and death from coronavirus disease 2019 (Covid-19), especially with the SARS-CoV-2 variants of concerns (VOC). It is unknown as to whether a history of Covid-19 prior to the vaccine and post-vaccine RBD-IgG levels are predictors of BNT162b2 vaccine effectiveness against VOC–δ in nursing home residents.

**Methods:** We analyzed the data from two NHs that faced a VOC-δ outbreak in July-August 2021. These NHs had suffered prior Covid-19 outbreaks in 2020 and 2021. In many of the residents, RBD-IgG levels were measured 6 weeks after the second vaccine dose, *i.e*. 3 to 5 months before the VOC-δ outbreak onset, and again during the outbreak (SARS-CoV-2 IgG II Quant assay, Abbott Diagnostics). We compared residents with vs without prior Covid-19 for (i) VOC-δ incidence, (ii) the correlation between post-vaccine RBD-IgG levels and VOC-δ incidence, and (iii) the time-related change in RBD-IgG levels.

**Results:** Among the 140 analyzed residents (58 to 101 years; 94 females, 46 men, mean age: 84.6 yr ± 9.5 yr), one resident among the 44 with prior Covid-19 before vaccination developed a VOC-δ infection during the outbreak (1.3%) vs 55 of the 96 without Covid-19 prior to vaccination (57.3 %)(p<0.0001). The median value for RBD-IgG 6 weeks after the vaccine and during the outbreak was higher in residents with prior Covid-19 (31,553 AU/mL and 22,880 AU/mL) than in those without (1,050 AU/mL and 260 AU/mL)(p<0.0001). In residents without Covid-19 prior to vaccination, post-vaccination RDB-IgG levels did not predict protection against VOC-δ infection.

**Conclusions:** In contrary to residents with prior SARS-CoV-2 infection, those without a history of Covid-19 before two BNT162b2 doses are not protected against VOC-δ infection and their RBD-Ig-G levels are low 3 to 5 months after vaccination. This suggests that a booster vaccine dose should be considered in this group of residents for a better protection against VOC-δ infection.

## Background

Nursing Home (NH) residents are at high risk of serious illness and death from coronavirus disease 2019 (Covid-19). Vaccination is safe and effective in adults, but is less documented in NH residents. ^1^ Outbreaks of SARS-CoV-2 ‘variants of concern’ (VOC), especially VOC–δ, have recently been described in NHs in whom most of the residents had received two doses of the BNT162b2 vaccine. ^2^

In March 2020, the French Occitanie region set up 22 Covid-19 support platforms to help NHs implement the Occitanie Health Agency recommendations (derived from the European Geriatric Medicine Society (EuGMS) guidance^3^) to prevent Covid-19 within their facilities. The Montpellier Covid-19 platform provides support to 122 NHs and over 6,000 residents ^4,5,6^ Specifically, as soon as a first case of positive RT-PCR is diagnosed in a resident, the support platform helps the NH (i) to implement infection, prevention and control (IPC) measures, (ii) to test all residents and staff members using real-time reverse-transcriptase polymerase chain reaction on nasopharyngeal swab test (RT-PCR), and (iii) to retest all RT-PCR-negative individuals weekly until no new cases are diagnosed for at least 14 days. ^4,6^ In collaboration with the platform, NHs may assess N-protein IgG levels in residents for whom prior Covid-19 is suspected to confirm diagnosis and post-vaccine S-protein (RBD)-IgG levels, in order to reinforce IPC measures in individuals with low post-vaccine antibody response.

We observed that a single dose of the BNT162b2 vaccine was sufficient in most NH residents with prior Covid-19 to obtain RBD-IgG levels of over 4,160 AU/mL, threshold associated with a high *in-vitro* neutralizing activity and equivalent to 433 BAU/mL. In contrast, 6 weeks after the second vaccine dose, 30% of NH residents without prior Covid-19 had RBD-IgG levels below 1,050 AU/mL, threshold predicting a neutralizing effect of the serum against SARS-CoV-2 *in vitro*. ^7,8,9^

The low immunogenicity of the vaccine in NH residents without prior Covid-19 and the fact that RBD-IgG produced by vaccinated and recovered individuals are less effective in binding and neutralizing *in vitro* VOC-β and δ than the “wild type” SARS-CoV-2 ^10,11^ could in part explain the occurrence of VOC-δ outbreaks in NHs in which most of the residents are vaccinated.

To determine whether vaccinated residents with Covid-19 before vaccination are more protected against the VOC-δ than those without, and whether residents with low post-vaccine RBD-IgG levels are at higher risk of incident VOC-δ, we analyzed the data from two NHs that faced a VOC-δ outbreak in July-August 2021. These facilities had already suffered prior Covid-19 outbreaks in 2020 and 2021. In many of the residents, RBD-IgG levels were measured 6 weeks after the second vaccine dose, i.e. 1,5 to 3,5 months before the VOC-δ outbreak onset, and again during the outbreak. We compared residents with vs without Covid-19 before vaccination for (i) VOC-δ incidence, (ii) the correlation between post-vaccine RBD-IgG levels and VOC-δ incidence, and (iii) the time-related change in RBD-IgG levels.

## Methods

### Settings and Participants

NH1 faced a Covid-19 outbreak in December 2020-January 2021. Residents received two BNT162b2 vaccine doses between March 11 and May 12, 2021, and the NH organized measurement of post-vaccine RBD-IgG levels 6 weeks after the second dose in most of the residents. NH1 faced a VOC-δ outbreak in July-August 2021 (first and last cases on July 23 and August 9, respectively). The NH organized RBD-IgG measurements in most of their residents between July 27 and July 29, 2021.

NH2 faced a Covid-19 outbreak in April-June 2020. Residents received two BNT162b2 vaccine doses between February 22 and March 5, 2021, and the NH organized measurement of post-vaccine RBD-IgG levels 6 weeks after the second dose in most of the residents. NH2 faced a VOC-δ outbreak in August 2021 (first and last cases on August 5 and 26, respectively). The NH organized RBD-IgG measurements in most of the residents on August 16 and September 2, 2021.

Residents were not tested (RT-PCR or blood sample) when the interest of the test was not considered by the NH coordinating practitioner as sufficient for the resident, or when the resident / family / relatives / legal representative refused the tests. Anonymized clinical and biological data were analyzed in accordance with the Montpellier University Hospital institutional review board approval (IRB-MTP _2021_04_202000534).

### Outcomes

**SARS-CoV-2 infection** was diagnosed using RT-PCR on nasopharyngeal swab (Allplex 2019-nCoV assay, Seegene). ^12^

**SARS-CoV-2 RBD IgG** levels were measured using the SARS-CoV-2 IgG II Quant assay (Abbott Diagnostics) 6 weeks after the second vaccine dose, and during the outbreak. Data for RT-PCR-positive residents were not analysed in the present work since Covid-19 infection can induce a rapid RBD-IgG synthesis.

## Results

### VOC-δ incidence rate according to Covid-19 history prior to vaccination

A total of 151 residents (58 to 101 years; 55 females, 26 men, mean age: 84.3 yr ± 9.5 yr in NH1, 49 females and 21 men, mean age: 85.8 yr ± 9.5 yr in NH2) were included. Among them, 140 (92.7%) had evaluable outcomes. Forty-four residents (31.4 %, 61 to 100 years, mean age: 84.6 yr ± 9.5yr) had prior Covid-19 before vaccination (positive RT-PCR in 2020 or 2021, or detectable SARS-CoV-2 nucleocapsid antibodies at the time of vaccination) (Table 1). One of them (1.3%) developed a VOC-δ infection during the outbreak.

**Table 1:** VOC-δ incidence (positive RT-PCR) in the 2 nursing home residents according to Covid-19 history prior to vaccination.

|  | NH1 | NH2 |
| --- | --- | --- |
| **Total number residents** | 81 | 70 |
| **Excluded** | 5 (4 Covid-19 between 2 vaccine doses) | 8 (2 non-vaccinated, 5 VOC-δ after vaccination, one undetermined prior Covid-19) |
| **Prior Covid-19 before vaccination, n (%)** | 38 (50.0%) | 6 (9.5%) |
| **Negative RT-PCR** | 38 (100%) | 5 (83.3%) |
| **Positive RT-PCR** | 0 (0%) | 1 (16.6%) |
| **No prior Covid-19 before vaccination, n (%)** | 38 (50.0%) | 56 (90.3%) |
| Negative RT-PCR | 24 (63.2%) | 40 (71.4%) |
| Positive RT-PCR | 14 (36.8%) | 26 (35.5%) |
| Death | 2 (5.3%) | 8 (12.9%) |

Among the 96 evaluable residents without Covid-19 prior to vaccination, 55 (57.3 %) developed a VOC-δ infection during the outbreak (59 to 101 years, mean age: 85.0 yr ± 8.9 yr).

### VOC-δ incidence rate according to post-vaccine RBD-IgG levels

RBD-IgG levels 6 weeks after the second vaccine dose were available in 84 residents (55.6 % of the whole sample, Table 2). All residents with prior Covid-19 before vaccination had post-vaccine RBD-IgG levels of over 1,050 AU/mL vs 4/25 (16%) residents without prior Covid-19 (p<0.0001). In residents without Covid-19 prior to vaccination, post-vaccination RBD-IgG levels did not predict the protection against VOC-δ infection.

**Table 2:**
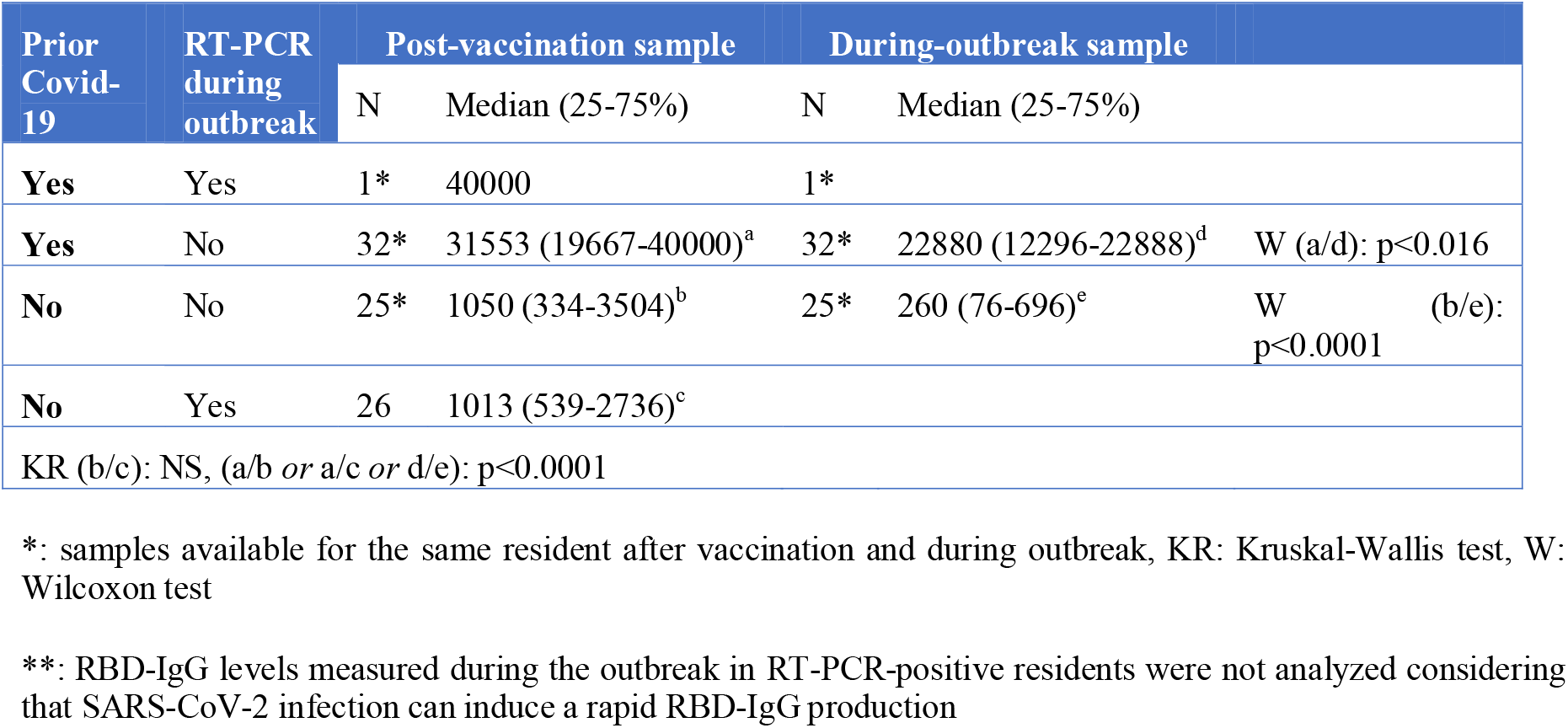
RDB-IgG levels measured 6 weeks after the second vaccine dose and during the outbreak according to Covid-19 history prior to vaccine and according to RT-PCR results during the outbreak **.

### Change in RBD-IgG levels between vaccination and the outbreak in RT-PCR negative residents

RBD IgG levels measured 6 weeks after the second vaccine dose and during the outbreak (3-5 months apart for NH1, and 5 to 6 months apart for NH2) were available in 57 residents who did not develop Covid-19 during the VOC-δ outbreak (32 with Covid-19 prior to the vaccine and 25 without Covid-19 prior to the vaccine, Table 2, Figure 1). There was a significant reduction of RBD-IgG levels in residents with Covid-19 prior to the vaccine (27.5% reduction) and in those without Covid-19 prior to the vaccine (69.6% reduction). In residents who did not develop Covid-19 during the outbreak, RBD-IgG levels were significantly higher in those with than in those without Covid-19 prior to the vaccine.

**Figure 1:**
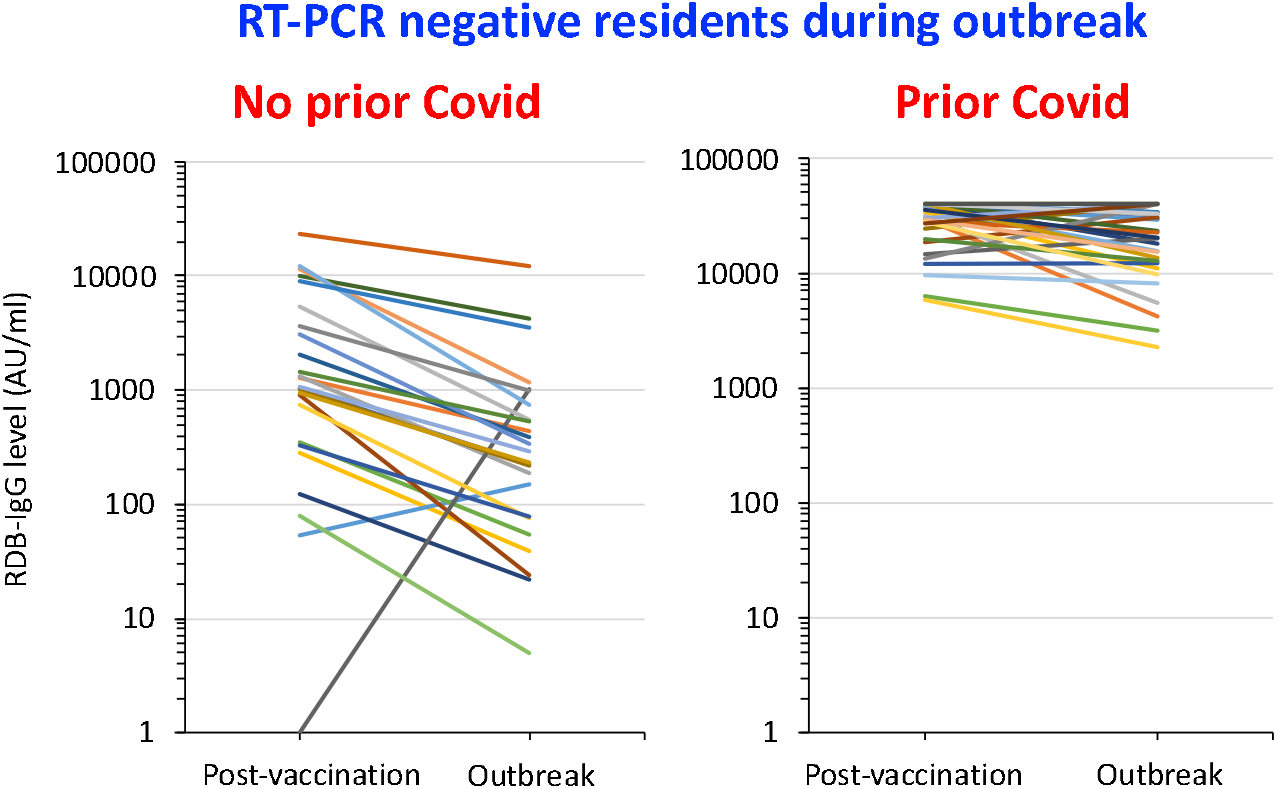
Change with time in RDB-IgG levels in residents with a negative RT-PCR during the outbreak: measures taken both 6 weeks after the second vaccine dose and during the outbreak.

## Discussion

The present study shows that residents with prior SARS-CoV-2 wild type infection 3 to 9 months before two BNT162b2 vaccine doses were almost totally protected from VOC-δ infection 3 to 5 months after their vaccination. At the same time, among fully vaccinated residents without Covid-19 prior to vaccine, most of them had low RBD-IgG levels and 57.3% of them developed a Covid-19 infection when they were faced with a VOC-δ outbreak in their NH.

As previously reported,^8^ two BNT162b2 vaccine doses induced a significantly higher post-vaccine immune response in residents with prior Covid-19 before vaccination than those without natural immunization. The analysis conducted in residents who do not develop Covid-19 shows that residents with Covid-19 prior to vaccination maintain a high level of RBD IgG at the time of VOC-δ outbreak, i.e. 1,5 to 3,5 months after vaccination, whereas most of the residents without Covid-19 before vaccination have low RBD-IgG levels at that time. RBD-IgG levels 6 weeks after the vaccine of residents without a Covid-19 history are comparable in those who developed or did not develop VOC-δ some months later. Thus, most of the residents without prior Covid-19 before vaccination had probably low RBD-IgG levels by the time the VOC-δ outbreak started in their NH. In this group, memory B cells and secondary immune response were probably most often not sufficient to prevent Covid-19. This result supports the hypothesis that a minimal post-vaccine RBD-IgG level may be required to block VOC-δ infection in NH residents, and that this minimal level is observed mainly in residents with Covid-19 prior to vaccination, and far less in those without. This result supports that a booster vaccine dose given less than 6 months after the second dose should be considered in NH residents without Covid-19 infection prior to vaccination in order better protect them against VOC-δ infection risk.

## Data Availability

All data referred to in the manuscript are available on request at the following address

## Acknowledgments

The authors would like to thank Anna Bedbrook, Fabienne Portejoie (MACVIA-France), and Eva Pons (Master Métiers de l’Enseignement, de l’Education et de la Formation; Education Nationale française, Lyon) for their editorial assistance; and Joy Martin, Marie-Béatrice Debru, Isabelle Vercruysse, Armelle Rochat, Secours Infirmiers, Véronique Vera, and Florence Biblocque (admissions office, Montpellier University Hospital) for material support. None of these contributors received any compensation for their help in carrying out the study.

